# Early-Life Environmental Exposures and Brain Structure in Schizophrenia

**DOI:** 10.64898/2026.10.22.26366215

**Authors:** Anja Stano, Juan F. Quinones, Djo Juliette Fischer, Sandra Weber, Leonie Klock, Ioanna Amaya, Robert C. Lorenz, Simone Kühn

## Abstract

**Background:** Schizophrenia is widely considered a neurodevelopmental disorder characterised by premorbid alterations in brain structure. Urban upbringing is a well-established environmental risk factor. However, urbanicity is often operationalised using broad population-based measures, leaving unclear which features of urban environments might affect the developing brain. This gap limits progress in understanding the pathways linking urban upbringing to schizophrenia, an increasingly important question as global urbanisation progresses. Cortical gyrification is especially informative in this context, as it is largely established before birth and remains relatively stable thereafter, offering a promising retrospective marker of early-life neurodevelopmental disturbances.

**Study Design:** In this case-control MRI study, we examined whether residential environmental exposures at birth were associated with adult brain structure in 51 individuals with schizophrenia or schizoaffective disorder and 88 healthy controls matched for age, sex and socioeconomic status. Birth addresses were linked to geospatial measures of air pollution, green space, built volume, built surface and population density.

**Study Results:** Higher air pollution at birth was associated with lower cortical gyrification in individuals with schizophrenia, but not in healthy controls, particularly in orbitofrontal and inferior frontal regions. No comparable associations were observed for grey matter volume, cortical thickness or surface area.

**Conclusions:** These findings suggest that air pollution at birth may be linked to hypogyria in schizophrenia and provide a potential bridge between epidemiological research on urbanicity and neurodevelopmental models of schizophrenia. In turn, this work provides a critical starting point for future work assessing how early urban environments become biologically embedded in schizophrenia risk.

## Introduction

Schizophrenia is a debilitating psychiatric disorder whose prevalence is projected to increase over the coming decades ^1^, thus highlighting the need to identify modifiable factors involved in its development. Decades of epidemiological research have consistently identified urbanicity as a significant risk factor ^2^. With urbanisation accelerating and around 68% of the world population projected to live in urban areas by 2050 ^3^, understanding the underlying mechanisms of this association is more important than ever. Previous research has made important progress by suggesting that the timing of urbanicity exposure matters, with early-life periods emerging as particularly relevant ^4^. This is consistent with the view of schizophrenia as a neurodevelopmental disorder, in which environmental disturbances during critical periods of early brain development may contribute to its emergence ^5–7^. Indeed, being born in an urban area has been consistently shown to increase the risk of schizophrenia ^8–13^ above and beyond genetic selection confounds ^10,12,14^. Proposed explanations for the association between urbanicity and schizophrenia involve psychosocial stressors clustered in cities and environmental hazards, such as air pollution ^2^.

The genuine effects of urbanicity have motivated questions about possible neurobiological pathways connecting urbanicity to mental health. Studies in healthy adults have related urban upbringing to reduced prefrontal grey matter volume (GMV) ^15,16^, altered activation of stress- and reward-related circuitry ^17,18^, reduced cortical thickness ^19^ and hippocampal volume ^20^, as well as less efficient prefrontal processing ^21^, which are all features that are implicated in schizophrenia ^22,23^. However, only very few studies have directly examined this question in patients ^24–26^. Beyond the small number of patient studies, this research line has been limited by how urbanicity has been defined. Most neuroimaging studies to date have relied on broad measures of urbanicity, often based on self-reported years spent in places of different population sizes during childhood ^27^. Cities, however, differ not only in population size, but also in terms of air pollution, noise, green space, and further characteristics of the built environment. This distinction is visible in epidemiological research, where exposure to air pollution has been linked to higher schizophrenia risk ^28,29^, whereas access to natural environments has been associated with lower risk ^30–32^.

At the same time, it is challenging to study how these environmental features relate to the brain in clinical samples. The exposure of interest often occurs early in life, while the brain is usually measured decades later, after illness onset. In the intervening years, the brain is influenced by maturation, illness processes, medication and later environmental exposures. Therefore, it is important to focus on brain metrics formed early in life that are less strongly affected by these intervening processes. Cortical folding, or gyrification, is a promising marker in this context. It refers to the formation of gyri and sulci that shape the cerebral cortex and has been proposed to reflect early neurodevelopmental processes involved in cortical expansion, neuronal migration and emerging cortical organisation ^33^. Gyrification undergoes its greatest development in utero ^34,35^, remains relatively stable after birth compared with other morphometric features ^36,37^ and appears sensitive to early-life environmental influences ^38,39^. Abnormal cortical folding has repeatedly been linked to schizophrenia ^40–42^. Nevertheless, only one previous study has examined urban birth and gyrification in schizophrenia, finding altered gyrification in patients but not healthy controls ^26^. However, this study also used population-based measures of urbanicity. Overall, it remains unclear which specific characteristics of urban environments may be biologically relevant for schizophrenia.

In this pre-registered case-control study (https://aspredicted.org/awMmvM.pdf), we aimed to address this gap in the literature by identifying features of the physical environment at birth associated with adult brain structure in individuals with schizophrenia and healthy controls. Gyrification was our main outcome of interest, given its relevance for early-life environmental influences. However, because fine-grained environmental features at birth have rarely been studied in relation to brain structure in schizophrenia, we additionally examined volumetric measures, cortical thickness and surface area.

## Methods

### Participants and procedure

All participants were recruited between July 2019 and July 2024. Before enrolment, potential participants were screened for MRI eligibility. Exclusion criteria included metallic implants, claustrophobia, tinnitus and pregnancy. After receiving a detailed explanation of the study procedures, all participants provided written informed consent and received financial compensation upon completion of the study. All participants first completed a brief sociodemographic interview, followed by additional computer-based questionnaires administered via Inquisit 6 ^43^, including information on residential history and socioeconomic status. Thereafter, participants underwent Magnetic Resonance Imaging (MRI) scanning. The study was approved by the local ethics committee of the University Medical Center Hamburg-Eppendorf, Germany (LPEK-006).

### Patients

A total of 51 patients with schizophrenia or schizoaffective disorder (SCZ) were recruited through the Department of Psychiatry and Psychotherapy at the Medical Center Hamburg-Eppendorf, Germany, and via advertisements. Diagnostic interviews were administered by seven trained raters employing a modified interview adapted from the German Mini International Neuropsychiatric Interview (MINI) ^44^. Participants were included if they met ICD-10 criteria for schizophrenia (F20) or schizoaffective disorder (F25) (ICD-10) ^45^ and had no severe neurological conditions (e.g. epilepsy, stroke).

### Healthy participants

88 healthy controls (HC) were recruited through public announcements and web-based advertisements. Psychiatric and neurological health was assessed using an adapted version of the Diagnostic Short Interview for Mental Disorders (MINI-DIPS) ^46,47^. HC were recruited to be comparable to the patient group in age, sex and socioeconomic background.

### Socioeconomic status

Socioeconomic status (SES) was assessed using an adaptation of the German Health Interview and Examination Survey for Adults (DEGS1) ^48^. Specifically, SES was quantified using a composite index based on education, occupation and household income. For education, highest school qualification and highest professional or university qualification were combined into one education score. For occupation, each reported job category was assigned a score reflecting its approximate occupational status, with higher scores for occupations requiring more training or responsibility. For income, the reported monthly household income category was replaced by an approximate euro value, divided by the household equivalence weight, and then converted into a seven-level income score. The education, occupation and income scores were summed to create the SES index, ranging from 3 to 21, with higher scores indicating higher socioeconomic status.

### Duration of antipsychotic medication

Duration of antipsychotic medication was assessed during the clinical interview by asking patients to report the number of years they had been taking antipsychotic medication.

### Environmental features

Residential environmental exposures were derived from participants’ birth addresses. Built surface, built volume and population density were obtained from the Global Human Settlement Layer (GHSL; 100 m spatial resolution raster files), available from 1975 to 2025 in five-year intervals. Built surface reflects the amount of land covered by built-up structures within each raster cell ^49^, whereas built volume reflects the total built-up volume in cubic metres and therefore captures the vertical dimension of the built environment ^50^. Population density represents the number of residents within each raster cell ^51^. Air pollution total emission data were obtained from the Emissions Database for Global Atmospheric Research (EDGAR) ^52^, available as yearly raster files at 0.1 by 0.1 degrees spatial resolution from 1970 to 2022. Pollutants included carbon monoxide (CO), nitrogen oxides (NOx), non-methane volatile organic compounds (NMVOC), sulphur dioxide (SO₂), particulate matter with aerodynamic diameter <10 μm (PM₁₀), particulate matter with aerodynamic diameter <2.5 μm (PM₂.₅), black carbon (BC) and organic carbon (OC). Green space was indexed using the Normalized Difference Vegetation Index (NDVI), obtained from the NASA Earthdata platform as monthly raster files at 0.0833 degrees spatial resolution, corresponding to approximately 9.3 km, and available from 1981 to 2022 ^53^. Monthly NDVI rasters were averaged to yearly values before extraction.

Birth address coordinates were geocoded and used to define circular buffers of 500 m and 1,000 m radius around each birth address. Environmental values were extracted in R (v.4.2) using the *terra* package ^54^. For each participant and environmental variable, raster values intersecting the buffer were weighted by the overlapping area between raster cell and buffer and subsequently summed across intersecting cells. For population density, built volume and built surface data, linear interpolation between available raster years was used to estimate values for the birth year. For air pollution emission data and NDVI, yearly rasters corresponding to the birth year were used. For birth years outside the temporal range of the available environmental data, the closest available raster year was used. The 500 m buffer was used as the primary exposure definition, with the 1,000 m buffer used in sensitivity analyses. Higher extracted values indicate greater environmental exposure at the address of birth. For sensitivity analyses adjusting for later environmental exposure, the same extraction procedure was applied to available yearly residential addresses from ages 1 to 15 years. Specifically, for each participant and environmental feature, mean exposure across these years was calculated and added as a covariate when testing the effect of exposure at birth.

### Magnetic resonance imaging data acquisition and processing

MRI data were acquired on a 3-T Siemens MAGNETOM Prisma scanner (Siemens Healthineers, Erlangen, Germany) equipped with a 64-channel head coil. At the beginning of each scanning session, structural images were obtained using a multi-parameter mapping (MPM) neuroimaging protocol ^55^ comprising three multi-echo spoiled gradient recalled echoes (SPGR) with proton density-weighted (PDw), T1-weighted (longitudinal relaxation time) and magnetisation transfer-weighted (MTw). Additional calibration sequences ^56,57^ were performed for B1+ correction. All scans were acquired with 1 mm isotropic resolution and a repetition time (TR) of 24.0 ms. Eight equidistant gradient echoes were collected for the PDw and T1w scans with echo times (TE) ranging from 2.3 ms to 18.4 ms (ΔTE = 2.3 ms), while six echoes were acquired for the MTw scan (TE = 2.3–13.8 ms, ΔTE = 2.3 ms). Flip angles were set to 6° for PDw and MTw, and 21° for T1w acquisition. MT-weighting was achieved by applying a 4 ms Gaussian-shaped off-resonance RF pulse (220° nominal flip angle, 2 kHz frequency offset) before excitation. Parallel imaging was employed with generalised autocalibrating partially parallel acquisitions (GRAPPA) acceleration factors of 3 (phase) × 1 (partition) and partial Fourier acquisition of 6/8 in both phase and partition directions. Total scan time was approximately 3 minutes and 46 seconds per multi-echo SPGR. Each multi-echo SPGR contrast was acquired twice within the same session to improve robustness against motion artefacts ^58^.

### MPM image reconstruction

MPM data were processed using the hMRI toolbox ^59^. PDw, T1w, MTw and B1+ images were used as input, together with a B0 field map to correct for magnetic inhomogeneity. This constructed a magnetisation transfer (MTsat) map, which has been shown to improve segmentation of subcortical areas, attributed to the high contrast-to-noise ratio ^60^, and reduced inter-site variability of grey matter segmentation compared to conventional T1w images ^55^. Therefore, this structural image was used as input for the CAT12 and Freesurfer analyses. All MRI scans were visually checked for quality control, and five participants had to be excluded from the CAT12 analyses due to image quality ratings (IQR) below 70.

### CAT12 regional GMV

Structural images were processed using the standalone version of the Computational Anatomy Toolbox (CAT12; https://neuro-jena.github.io/cat/) on a high-performance computing cluster. The cross-sectional segmentation pipeline included image denoising, spatial registration, correction for intensity inhomogeneity, skull stripping, tissue classification and partial-volume estimation. GMV was estimated from the segmented tissue classes. Regional GMV was extracted using the CAT12 Hammers atlas ^61^. The ROI analyses included 95 grey matter regions, comprising 46 left-hemispheric regions, 46 right-hemispheric regions, as well as three non-lateralised regions, corresponding to the third ventricle, brainstem excluding substantia nigra and corpus callosum.

### FreeSurfer cortical, subcortical and gyrification measures

Cortical reconstruction was performed using the FreeSurfer version 8.1.0 recon-all pipeline. Global brain volume measures, estimated total intracranial volume (eTIV) and subcortical structure volumes were extracted from the FreeSurfer aseg.stats files. The cortex was parcellated according to the Desikan-Killiany atlas, comprising 34 cortical regions per hemisphere and 68 cortical regions in total. For each cortical region, cortical thickness and surface area were extracted from the FreeSurfer aparc.stats files. Local gyrification index (LGI) was computed using the FreeSurfer recon-all -localGI procedure. In this procedure, LGI is first calculated at each vertex of the reconstructed pial surface as the ratio between the amount of cortex buried within the sulci and the amount of cortex visible on the outer surface within a local circular region. Regional LGI values were then obtained by averaging vertex-wise LGI values within each Desikan-Killiany parcel, resulting in one LGI value per cortical region.

### Statistical analysis

All statistical analyses were performed using R Statistical Software (v.4.5.3). Between-group differences in MRI-derived morphometric measures were tested using linear regression models adjusted for age, sex and pipeline-specific total intracranial volume, using eTIV for FreeSurfer measures and TIV for CAT12 measures. Associations between residential environmental exposures at birth and MRI-derived measures were tested using separate linear models for each exposure-outcome pair. Environmental exposure variables were standardised prior to modelling. Main-effect and interaction-effect models were adjusted for diagnostic group, age, sex, socioeconomic status, intracranial volume and duration in years of antipsychotic medication. Interaction models additionally included an exposure × diagnostic group term. To examine whether significant findings reflected exposure at birth rather than environmental exposure during later upbringing, significant effects were rerun with additional adjustment for later exposure to the same environmental feature. Specifically, for each participant, mean exposure to the same environmental feature from ages 1 to 15 years was calculated using the same buffer size and added to the respective model as an additional covariate.

To account for multiple comparisons, p-values from the exposure-brain regression models were corrected using the Benjamini-Hochberg false discovery rate (FDR) procedure ^62^ separately within each morphometric domain (global measures, cortical thickness, surface area and LGI) and separately for main-effect and interaction models. For between-group differences, FDR correction was applied across all included brain-structure measures and, separately, across all included environmental exposure measures. Healthy controls were used as the reference group, such that negative group coefficients indicate lower values in the schizophrenia group.

Outliers and influential observations were assessed as follows. For FDR-significant regression estimates, potential influence of individual observations was assessed by refitting each model after omitting one participant at a time and comparing the interaction estimates with those from the full sample ^63^. Morphometric outliers were identified using Tukey’s interquartile range (IQR) rule ^64^. Values that were 1.5 x IQR beyond the first or third quartile were set to missing for the respective measure, and the analyses were repeated. Outliers were defined separately within each brain measure as well as within each case *vs.* control group to avoid removing values due to true between-group differences.

### Missing data

Antipsychotic medication duration was missing for one schizophrenia participant with documented antipsychotic treatment. For this participant, medication duration was estimated as the midpoint between zero and the maximum possible illness duration, defined as the time between first psychotic episode and MRI assessment. Missing values in socioeconomic status (*N*_HC_ = 6, *N*_SCZ_ = 1) were imputed using predictive mean matching as implemented in the *mice* package in R. Five imputed datasets were generated, and the first completed dataset was used for the analyses. To assess whether the findings depended on the selected dataset, estimates were also pooled across all five imputed datasets and resulted in no meaningful differences. Imputation was based on age, sex, diagnostic group, estimated total intracranial volume and duration of antipsychotic medication.

## Results

### Sample

The sample included 51 patients and 88 healthy controls. Because MRI availability differed by processing pipeline and environmental exposure data were not available for all participants, complete-case sample sizes varied across analyses. FreeSurfer analyses included up to 114 participants, whereas CAT12 analyses included up to 104 participants after additional exclusions for low image-quality related interquartile range values. Table 1 summarises participant characteristics, including availability of birth addresses and MRI data.

**Table 1.** Sample characteristics.

| Characteristic | HC | SCZ |
| --- | --- | --- |
| *N* | 88 | 51 |
| Age, years | 35.3 ± 9.3 | 39.1 ± 10.9 |
| Female sex, n (%) | 44 (50%) | 28 (54.9%) |
| SES index | 11.7 ± 3.3 | 9.1 ± 2.6 |
| Handedness, right-handed (%) | 94.0% | 84.3% |
| Birth address available, n (%) | 82 (93.2%) | 45 (88.2%) |
| MRI data available, n (%) | 79 (89.8%) | 46 (90.2%) |
| Age at first episode, years | – | 24.3 ± 8.3 |
| Number of episodes, median [IQR] | – | 3 [2.0-5.6] |
| Number of inpatient treatments, median [IQR] | – | 4.5 [2-7.8] |
| Years of antipsychotic medication, median [IQR] | – | 8 [1-10] |
| ICD-10 diagnosis F20 (%) | – | 74.5% |
| ICD- 10 diagnosis F25 (%) | – | 25.5% |

*Note.* Values are shown as mean ± SD unless otherwise specified. SES = socioeconomic status; the SES index ranged from 3 to 21, with higher values indicating higher socioeconomic status. IQR = interquartile range.

### Intercorrelations among environmental features at birth

Environmental exposures at birth were strongly interrelated (Figure 1). Air pollutants, population density and built environment measures were moderately to strongly positively correlated with one another, whereas green space showed moderate to strong negative correlations with these features. The complete correlation matrix with exact Pearson coefficients is provided in Supplementary Figure S1.

**Figure 1.**
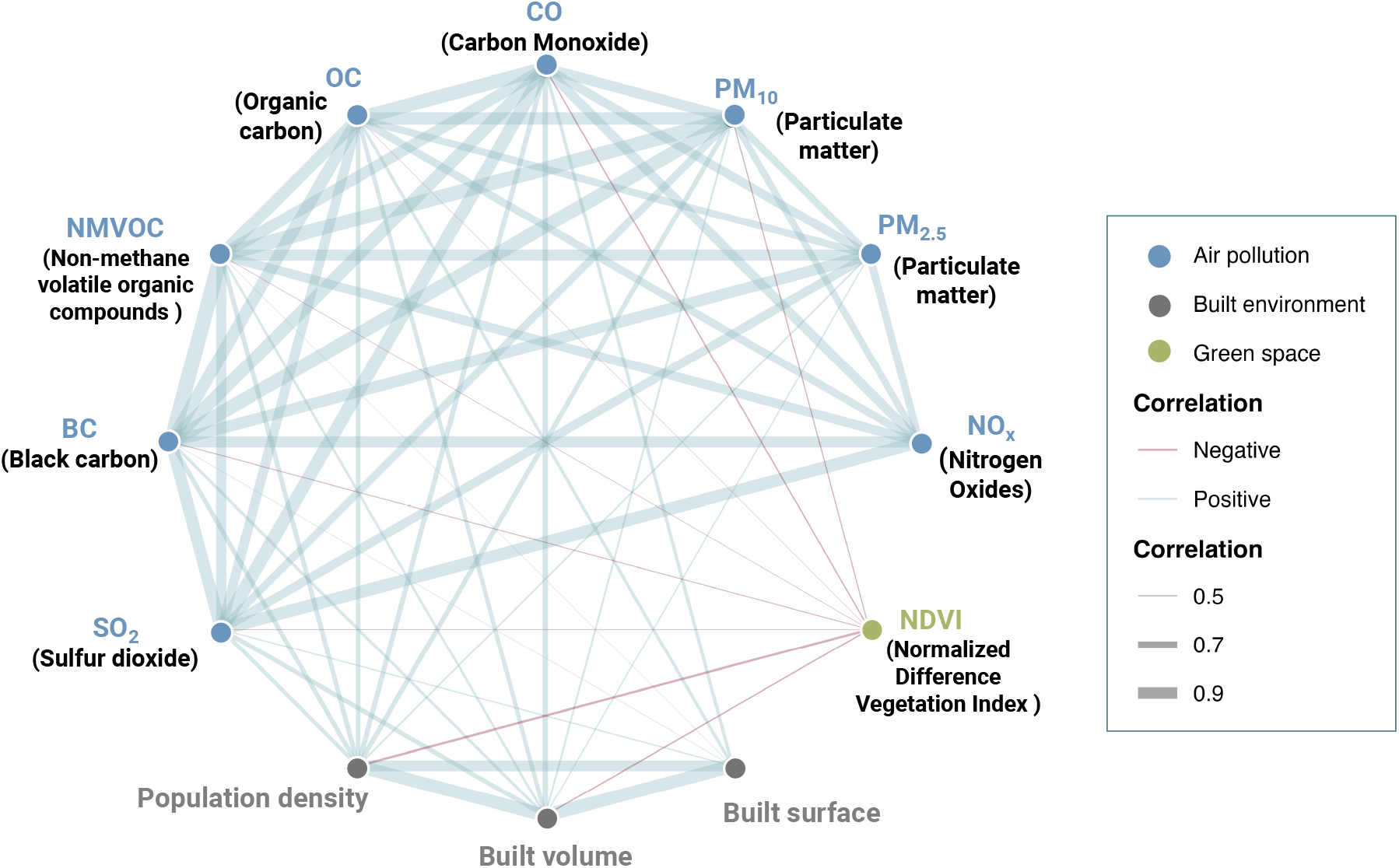
Correlation network of Pearson correlations between residential environmental exposures at birth, extracted using the 500 m buffer. Displayed correlations ranged from r = -.34 to r = .99, with darker and thicker edges indicating stronger correlations.

### Between-group differences in environmental exposures at birth

Compared with healthy controls, individuals with schizophrenia showed higher exposure values at birth across several environmental measures at the 500 m buffer, including BC, built volume, built surface, NMVOC, CO, NO_x_, PM_10_, PM_2.5_ and population density (all *p*FDR ≤ .036; Figure 2; Supplementary Table S1). Between-group differences in green space, OC and SO_2_ were marginal but were not significantly different from zero, although effects were in the expected direction with lower green space and higher OC and SO_2_ in schizophrenia. Results were consistent at the 1,000 m buffer, with effects in the same direction and of comparable magnitude (Supplementary Table S1).

**Figure 2.**
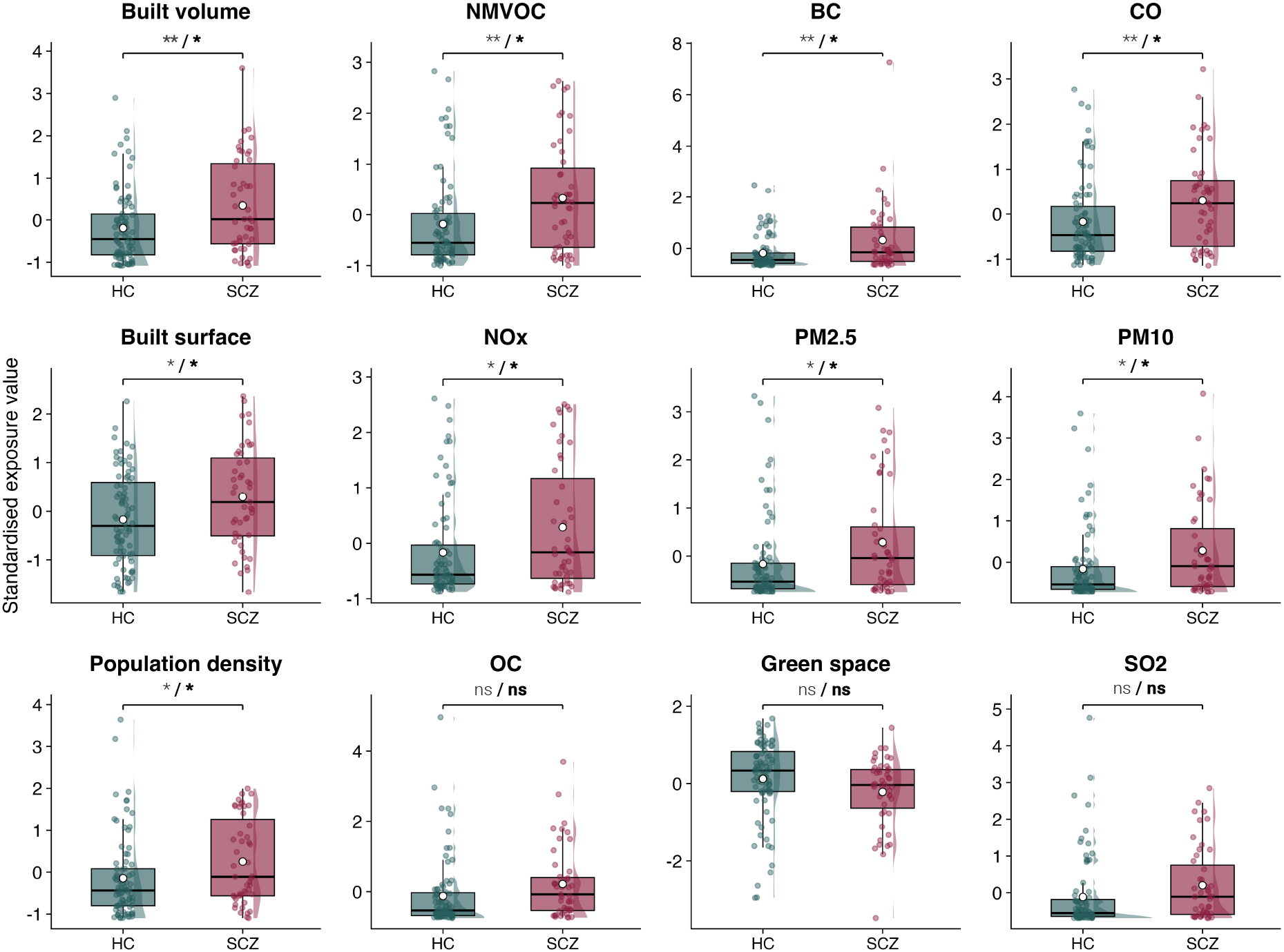
Between-group differences in residential environmental exposures at birth. Standardised residential environmental exposures at birth by diagnostic group, based on the 500-m buffer (*N* = 127). HC indicates healthy controls; SCZ, participants with schizophrenia. Points represent individual participants, boxes show medians and interquartile ranges, half-violin densities show the distributions, and white circles indicate group means. Exposure values were standardised within environmental variable. Brackets indicate diagnostic-group comparisons, with the first asterisk (*) referring to the uncorrected p-value and the second to the FDR-corrected p-value: * *p* < .05; ** *p* < .01; ns = not significant.

### Between-group differences in brain structure

Among the 125 participants with MRI data (HC = 79, SCZ = 46), between-group differences were observed mainly in global, cortical, and subcortical volume measures after FDR correction across all tested brain structural measures. Compared with healthy controls, individuals with schizophrenia showed lower total cortical volume (β = −20,311.13; *p* < .001; *p*FDR = .007), total GMV (β = −18,580.06; *p* = .002; *p*FDR = .020), and brain volume excluding ventricles (β = −26,362.89; *p* = .002; *p*FDR = .020). Subcortically, individuals with schizophrenia showed larger right pallidum (β = 137.08; *p* < .001; *p*FDR < .001), left pallidum (β = 122.56; *p* < .001; *p*FDR = .001), right putamen (β = 272.67; *p* = .002; *p*FDR = .014), right accumbens area (β = 42.69; *p* = .002; *p*FDR = .020), left putamen (β = 247.91; *p* = .003; *p*FDR = .026), and left caudate volumes (β = 220.06; *p* = .003; *p*FDR = .028). For cortical measures, schizophrenia was associated with lower surface area in the left caudal anterior cingulate cortex (β = −90.61; *p* < .001; *p*FDR = .014) and lower LGI in the left postcentral cortex (β = −0.120; *p* = .001; *p*FDR = .040), right postcentral cortex (β = −0.121; *p* = .001; *p*FDR = .040), left precentral cortex (β = −0.117; *p* = .002; *p*FDR = .041), and right rostral middle frontal cortex (β = −0.087; *p* = .002; *p*FDR = .041). The association with right precentral LGI was trend-level (β = −0.106; *p* = .004; *p*FDR = .052). No associations involving cortical thickness or CAT12-derived regional GMV survived FDR correction. Complete regression results are provided in Supplementary Table S8.

### GMV, cortical thickness, surface area and environmental exposures

No associations between residential environmental exposures at birth and FreeSurfer-derived measures of global or subcortical volume, hemispheric mean cortical thickness, regional cortical thickness, or regional surface area survived FDR correction in either the 500 m or 1000 m buffer analyses. In CAT12 regional GMV analyses, 500 m urbanicity-related exposures were associated with lower GMV in the left anterior insula. Specifically, population density at birth was negatively associated with GMV in the left anterior inferior insula (β = −0.293; 95% CI, −0.461 to −0.124; *p* < .001; *p*FDR = .041) and left anterior short insula (β = −0.282; 95% CI, −0.441 to −0.122; *p* < .001; *p*FDR = .041). Built volume was also, marginally, negatively associated with GMV in the left anterior inferior insula (β = −0.295; 95% CI, −0.458 to −0.133; *p* < .001; *p*FDR = .048) and left anterior short insula (β = −0.291; 95% CI, −0.464 to −0.118; *p* = .001; *p*FDR = .057), as illustrated in Figure 3. The corresponding 1000 m associations did not survive FDR correction at *p*FDR < .05, although built volume showed trend-level associations with the same left anterior insula regions. After additional adjustment for exposure to built volume and population density during the first 15 years of life, the GMV associations were no longer significant. Finally, no exposure × diagnostic group interactions survived FDR correction for CAT12 regional GMV or FreeSurfer-derived cortical/subcortical morphometric outcomes after adjustment for age, sex, socioeconomic status, total intracranial volume and duration of antipsychotic medication. Complete CAT12 GMV results are reported in Supplementary Tables S2 (500 m) and S3 (1000 m), and complete FreeSurfer results in Supplementary Tables S4 (500 m) and S5 (1000 m).

**Figure 3.**
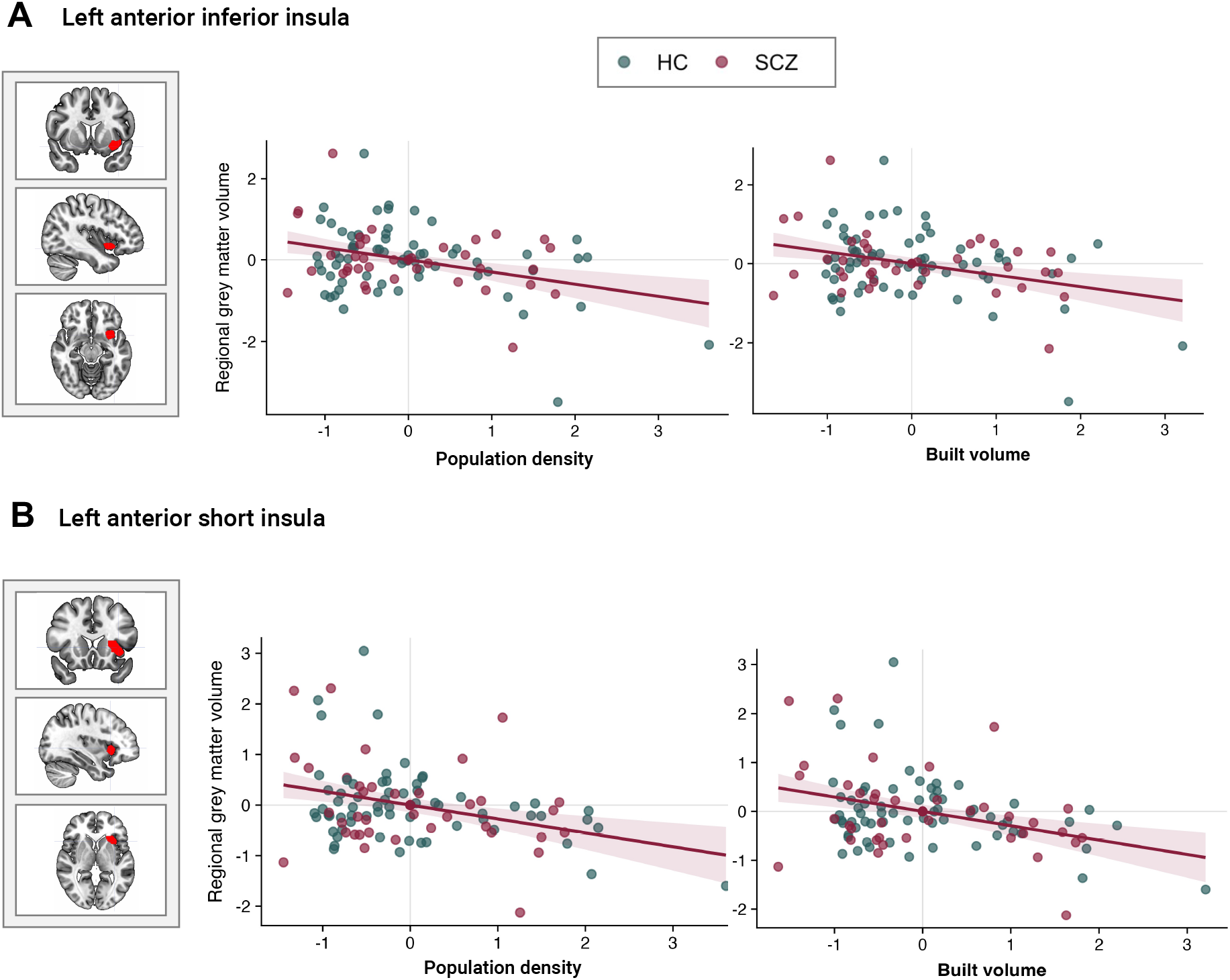
Population density and built volume at birth are associated with lower left anterior insular GMV. Associations between residential environmental exposures at birth within the 500-m buffer and CAT12-derived regional GMV in two left insular subregions (N = 104; HC = 65, SCZ = 39). HC indicates healthy controls; SCZ, participants with schizophrenia. (A) Left anterior inferior insula. (B) Left anterior short insula. Points represent individual participants, colored by diagnostic group. Lines show model-predicted associations with 95% confidence intervals after adjustment for diagnostic group, age, sex, socioeconomic status, total intracranial volume, and duration of antipsychotic medication. Population density survived FDR correction in both regions. Built volume survived FDR correction in the left anterior short insula and showed a trend-level association in the left anterior inferior insula.

### Cortical gyrification and environmental exposures

When the whole sample was analysed, no main effects of residential environmental exposures at birth on LGI survived FDR correction. However, the association between air pollution emissions at birth and LGI differed by diagnostic group. At the 500 m buffer, FDR-corrected significant interaction effects were found for OC, CO, PM10 and PM2.5, localising to orbitofrontal and inferior frontal LGI regions (Figure 4). Across pollutants, these effects involved right pars orbitalis, right lateral orbitofrontal cortex, right pars triangularis, right pars opercularis and left lateral orbitofrontal cortex. All interaction estimates were negative, indicating that higher air pollution was associated with lower LGI in schizophrenia relative to healthy controls. OC showed the largest negative interaction estimates, including the right pars orbitalis (interaction β = −0.698; 95% CI, −1.036 to −0.359; *p* < .001; *p*FDR = .006), right pars triangularis (interaction β = −0.627; 95% CI, −0.958 to −0.295; *p* < .001; *p*FDR = .009), right lateral orbitofrontal cortex (interaction β = −0.597; 95% CI, −0.919 to −0.274; *p* < .001; *p*FDR = .009), right pars opercularis (interaction β = −0.570; 95% CI, −0.894 to −0.245; *p* < .001; *p*FDR = .012), and left lateral orbitofrontal cortex (interaction β = −0.557; 95% CI, −0.887 to −0.228; *p* = .001; *p*FDR = .015). Comparable effects were observed for CO, PM10 and PM2.5 in overlapping orbitofrontal and inferior frontal regions (Supplementary Table S6). Sensitivity analyses using the 1000 m buffer showed a similar spatial pattern, with effects again localising primarily to orbitofrontal and inferior frontal regions. At this larger buffer, additional FDR-significant interactions were observed for NMVOC and BC, and PM_2.5_ additionally showed an interaction in the right insula. These results are reported in Supplementary Table S7.

**Figure 4.**
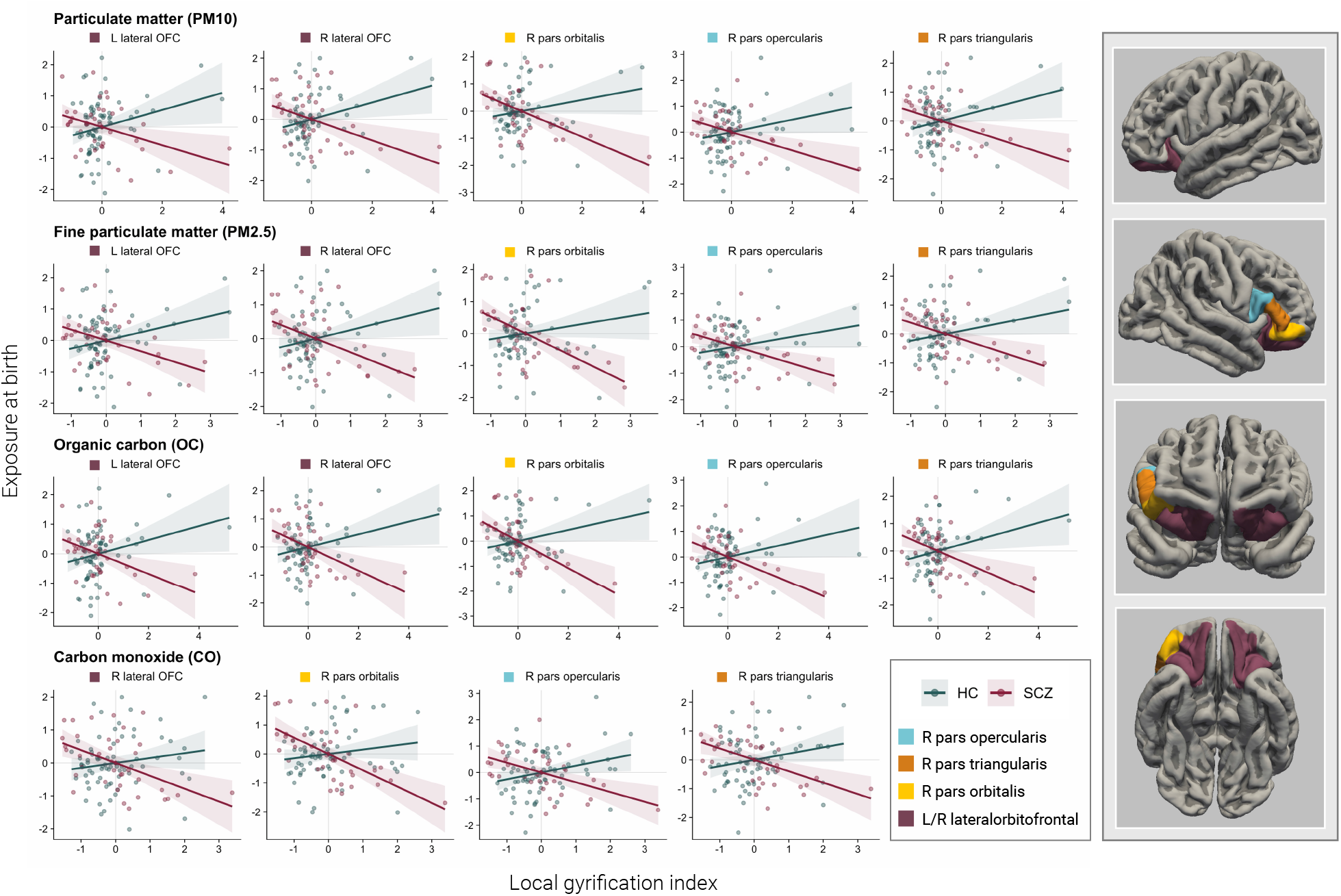
Air pollution exposure at birth and cortical gyrification in schizophrenia and healthy controls. Scatterplots show FDR-significant exposure x diagnosis interactions between individual air pollutants at birth within the 500 m buffer and LGI. Panels show partial associations after residualising both LGI and environmental exposure for age, sex, socioeconomic status, estimated total intracranial volume and duration of antipsychotic medication. Points represent individual participants and regression lines are shown separately for healthy controls and individuals with schizophrenia, with shaded 95% confidence intervals. Higher exposure was associated with lower LGI in individuals with schizophrenia, but not in healthy controls, primarily in orbitofrontal and inferior frontal regions. HC indicates healthy controls; SCZ, participants with schizophrenia.

Group-stratified models were used to describe the direction of the group-specific associations. In schizophrenia, the slopes were negative across the significant interaction findings, whereas no associations survived FDR correction in healthy controls. In schizophrenia, FDR-corrected associations were observed for CO and OC in right pars orbitalis LGI. Specifically, CO was negatively associated with right pars orbitalis LGI (β = −0.524; 95% CI, −0.761 to −0.287; *p* < .001; *p*FDR = .006), as was OC (β = −0.500; 95% CI, −0.769 to −0.231; *p* < .001; *p*FDR = .042). Additional associations within the schizophrenia group with OC, CO and PM10 in overlapping orbitofrontal and inferior frontal regions were below p < .01 uncorrected, but did not survive FDR correction at *p*FDR < .05. To examine whether the interaction findings were independent of environmental exposures later in childhood, significant LGI interaction models were additionally adjusted for mean exposure to the same environmental feature during upbringing from ages 1 to 15 years. The interaction estimates remained largely unchanged. All respective regression coefficients are reported in Supplementary Table S6.

### Sensitivity analyses

First, to assess whether the FDR-significant LGI interactions were driven by individual potentially influential observations, each model was refitted after omitting one participant at a time. All interaction estimates remained negative and statistically significant after exclusion of any individual participant, indicating that the findings were not dependent on outliers.

Second, we examined whether the findings were affected by outlying morphometric values using the IQR rule. Overall, results were comparable to the primary analyses. The LGI exposure × diagnosis interactions remained robust and showed the same regional pattern, with strongest effects in orbitofrontal and inferior frontal regions. No additional volume or surface-area associations survived FDR correction. In cortical thickness analyses, NDVI showed additional trend-level positive associations after outlier removal, most prominently in left middle temporal cortex (β = 0.293, *p* = .0018, *p*FDR = .069) and right pars triangularis (β = 0.273, *p* = .0020, *p*FDR = .069). Further trend-level associations were observed in left insula (β = 0.247, *p* = .0054, *p*FDR = .092) and right pars orbitalis (β = 0.273, *p* = .0050, *p*FDR = .092).

## Discussion

In this study, we sought to better understand the mechanisms behind the longstanding epidemiological finding that early-life urbanicity increases risk for schizophrenia. To do so, we extended beyond previous literature that primarily relied on broad definitions of urbanicity by instead focusing on specific physical environmental features at birth. We found that higher air pollution emissions at birth were associated with lower adult cortical gyrification in individuals with schizophrenia, but not in healthy controls. These associations survived correction for multiple comparisons and adjustment for age, sex, socioeconomic status, total intracranial volume and duration of antipsychotic medication. Further adjustment for exposure during later upbringing did not affect the results, suggesting that conditions around birth, potentially during the pre- and perinatal period, may be particularly relevant.

### Physical Environment at Birth and Adult Brain Structure

Among the gyrification findings, the strongest associations were observed for OC, CO, PM_10_ and PM_2.5_ and were located mainly in ventral frontal regions, including lateral orbitofrontal cortex and inferior frontal gyrus. These regions overlap with areas implicated in schizophrenia ^65–67^, offering a possible indication of how early-life urbanicity may relate to schizophrenia risk through structural brain alterations. Our findings extend the only previous study of urbanicity and cortical folding in schizophrenia, which also reported altered gyrification associated with urban birth ^26^. Similar to that study, the associations in our data were specific to the schizophrenia group and absent in healthy controls. This is also consistent with a previous study in healthy participants, where urban upbringing was not associated with gyrification ^19^. The restriction of the findings to schizophrenia patients is consistent with evidence that genetic vulnerability may modify associations between air pollution and outcomes related to psychosis. In a twin study, the association between air pollution and psychotic experiences depended partly on genetic vulnerability (Stano et al., *submitted*). Similarly, a prospective study reported that genetic susceptibility modified the association between long term air pollution exposure and schizophrenia risk ^68^. Recent evidence has also linked air pollution to adolescent brain outcomes, with some effects varying by schizophrenia genetic liability ^69^. Air pollution may therefore be more consequential in genetically vulnerable individuals.

### Developmental Timing of Environmental Effects

The specificity of the findings to cortical folding is plausible from a developmental perspective. Schizophrenia is widely considered a neurodevelopmental disorder, in which disturbances during early brain development may contribute to later vulnerability to psychosis ^5,70^. Environmental exposures around birth are therefore particularly relevant, since they occur during a critical period of neurodevelopment ^71^. Gyrification is an important marker in this context, because it is largely established in prenatal life and remains relatively stable across the lifespan ^35,72^. Twin studies further suggest that gyrification in the regions identified in this study is only partly heritable and shows substantial non-shared environmental variance ^39^, making environmental associations in these regions plausible.

By contrast, birth exposures were not robustly associated with global or regional GMV, cortical thickness or surface area. Some associations between regional GMV and the built environment and population density were initially observed, but they were no longer significant after adjustment for exposures during later upbringing. GMV and cortical thickness are more dynamic structural measures that continue to change with maturation, ageing and environmental experience ^34^, and may therefore be less reflective of the environment at birth. In schizophrenia, they may additionally be influenced by illness-related processes and treatment exposure, including progressive cortical thinning around psychosis onset ^73^ and structural effects of antipsychotic medication ^74^. Surface area, in turn, is considered one of the most heritable cortical measures ^75^, potentially leaving less variance attributable to environmental conditions at birth. Further, surface area also changes across development and adulthood ^76,77^. In this same dataset, cumulative exposure to population density across the first 15 years of life was associated with surface area (Quinones et al., *in preparation*), suggesting that surface area may reflect environmental influences accumulated across development rather than conditions at birth.

### Potential Biological Mechanisms

Air pollution could plausibly interfere with early-life neurodevelopmental processes through several biological pathways. Inhaled particles such as black carbon have been detected on the fetal side of the human placenta, suggesting that environmental particles can reach the fetal compartment ^78^. During pregnancy, air pollution may also affect fetal development indirectly through placental dysfunction, oxidative stress and epigenetic changes ^79^. These processes are relevant for the present findings because intrauterine conditions such as hypoxia, inflammation and altered placental function can influence cortical maturation and folding trajectories ^38^. Importantly, several biological responses to air pollution overlap with processes implicated in schizophrenia, including dysregulation of the hypothalamic– pituitary–adrenal (HPA) axis, neuroinflammatory signalling and oxidative stress ^80–82^. The neural diathesis-stress model of schizophrenia further proposes that HPA axis activation in response to stress can amplify underlying dopaminergic abnormalities associated with psychosis ^81,83^. Air pollution related biological responses may therefore have greater neurodevelopmental consequences in individuals with underlying vulnerability to psychosis, which may help explain why the association with gyrification was observed in the schizophrenia group but not in healthy controls.

### Strengths

To our knowledge, this is the first case-control neuroimaging study in schizophrenia to combine patient data with geospatial information on specific features of the physical environment at birth. This is important as most neuroimaging studies focusing on urbanicity effects have been conducted in healthy samples and have relied on population-based measures. Yet urbanicity is a mixture of correlated environmental features that may differ in their biological relevance and developmental timing. By differentiating between measures of air pollution, green space, population density and the built environment, we could show that the gyrification findings were driven by air pollution but not by the other environmental features. Such specificity makes the finding more interpretable and provides a clearer starting point for future studies of biological mechanisms. The decomposition of urbanicity was also important when accounting for exposures during later upbringing. Previous studies based on population-based urbanicity suggested that urbanicity during the first 15 years of life fully explains associations initially attributed to urbanicity at birth ^84^. However, such broad measures cannot show whether the same conclusion applies to all features of urban environments. In our study, the association between air pollution at birth and gyrification remained robust after adjustment for later exposure to the same environmental feature. This suggests that conclusions about the timing of urbanicity effects may depend on which component of the urban environment is being measured, with air pollution likely playing a distinct role in the perinatal period.

### Limitations

A primary limitation of this work is its cross-sectional design, with MRI measurements acquired in adulthood. We therefore cannot infer causality or reconstruct the developmental trajectory from conditions at birth to adult brain structure. Social drift after illness onset is unlikely to explain the findings, as environmental measures were derived from addresses at birth and thus preceded illness onset. However, intergenerational selection into urban environments remains possible. Families with greater schizophrenia liability may be more likely to live in polluted areas before a child’s birth. We could not test this possibility directly because genetic data were unavailable. However, previous studies have shown that the association between urban upbringing and schizophrenia risk persists after adjustment for polygenic risk scores ^10,14,85,86^, suggesting that urbanicity may be associated with schizophrenia risk beyond inherited liability. Nevertheless, future studies should incorporate genetic measures to determine whether the association remains after accounting for inherited liability.

Second, the sample size was modest and was further reduced by MRI and environmental data availability. Environmental exposure assessment depended on retrospective residential history, which can be difficult to reconstruct several decades later, particularly in clinical samples. Therefore, missing or incomplete address histories may have reduced statistical power and limited the generalisability of the findings.

Finally, schizophrenia is clinically heterogeneous, and our patient sample included individuals with schizophrenia (F20) as well as schizoaffective disorder (F25) according to ICD-10 diagnoses, limiting our ability to determine whether the observed associations apply equally across diagnostic subgroups or clinical profiles. Replication in larger independent samples integrating symptom profiles will therefore be essential to test whether specific features of urbanicity, such as air pollution, are consistently related to cortical folding.

## Conclusion

In conclusion, this study suggests that air pollution at birth is a key environmental feature through which urbanicity may become biologically relevant for schizophrenia. Specifically, we found that air pollution was linked to cortical gyrification, primarily in ventral frontal cortical regions, in individuals with schizophrenia but not healthy controls. Given that gyrification is primarily established in the pre- and perinatal period, the findings highlight that air pollution around birth may be particularly relevant in the context of schizophrenia vulnerability. Importantly, as air pollution is a modifiable environmental risk factor, these findings can be used to inform prevention strategies to help mitigate the rising trend in schizophrenia incidence. These findings should nevertheless be interpreted cautiously, as intergenerational selection into urban environments cannot be fully excluded. Future studies in larger clinical samples should replicate these findings and incorporate genetic liability to determine whether air pollution at birth remains associated with neurodevelopmental brain outcomes after accounting for inherited risk.

## Data Availability

Data generated in the present study are available from the corresponding author upon reasonable request, except for residential address information, which cannot be shared because of participant privacy and data-protection restrictions.

## Acknowledgements

During the work on her dissertation, A.S. was a pre-doctoral fellow of the International Max Planck Research School on the Life Course (IMPRS LIFE, www.imprs-life.mpg.de; participating institutions: Max Planck Institute for Human Development, Freie Universität Berlin, Humboldt-Universität zu Berlin, Technische Universität Berlin, University of Michigan, University of Virginia, and University of Zurich).

## Author Contributions

AS: Conceptualisation, Methodology, Formal analysis, Visualisation, Writing – original draft. JFQ: Conceptualisation, Methodology, Writing – review and editing. DJF: Methodology, Writing – review and editing. SW: Data collection, Writing – review and editing. LK: Data collection, Writing – review and editing. IA: Writing – review and editing. RCL: Processing of multi-parameter mapping (MPM) data, Writing – review and editing. SK: Conceptualisation, Supervision, Writing – review and editing. All authors read and approved the final manuscript.

